# Validation of Remote Testing using BrainCheck, a Computerized Neurocognitive Test

**DOI:** 10.1101/2020.06.01.20119289

**Authors:** Siao Ye, Bin Huang, Kevin Sun, Huy Phi, Reza Hosseini Ghomi

## Abstract

Remote computerized neurocognitive testing (CNT) is a promising solution to have these assessments more accessible to a population facing a global pandemic and increased aging. BrainCheck (BC) is a CNT software available on iPhone, iPad, and computer browser, designed to fit the need for remote testing. Consistent measures across these varying platforms are necessary to ensure users have consistent cognitive assessments and results. We aimed to assess BC across all administration platforms and interactions to observe any differences in assessment performance. 75 participants were enrolled in the study and were divided into two groups: participants who took BC across multiple platforms and participants who took BC in both an administered and self-administered fashion. Here we found Stroop, Digit Symbol, and Trail A/B had significantly different performance across the platforms, while Flanker, Coordination, Matrix, Immediate and Delayed Recognitions did not. Also, we found that the test metrics did not show significant differences in performance between being administered and self-administering the test. We did observe quicker completion times during the second instance of the test when taken in quick succession (within a day apart, which would not be a typical clinical pattern) and despite this, composite scores did not change reflecting the resilience of BC to practice effects. In conclusion, our results demonstrate BC may be a robust, self-administered CNT solution with an appropriate adjustment for the platform used.

## Introduction

The significant advantages of computerized neurocognitive testing (CNT) are clear: efficiency, cost, and accessibility. While the capability of remote CNT was present prior to the time of this study, the demand wasn’t as high. However, with the World Health Organization declaring COVID-19 as a global pandemic on March,11, 2020 [1], the initiation of interventions to reduce the spread of this disease limited non-essential medical care. To address this, CMS guidelines started to support reimbursement for cognitive testing via telehealth during the pandemic and with elderly patients being most vulnerable and many living in facilities not allowing off site trips, the immediate need for remote cognitive testing arised.

Supporting remote CNT is not only an urgent solution to meet current challenges but is also a long-term solution for the substantial increase of dementia patients in the near future, predicted to reach 7.1 million by the year of 2025 [2]. The digitized and remotely accessible CNT will be critical to lower the barriers for early detection, diagnosis, and management of cognitive disorders including: Alzheimer’s Dementia (AD), mild cognitive impairment (MCI), mild traumatic brain injury (mTBI), and any change in cognition due to underlying neurodegenerative disease or otherwise. Video-administered screening has already been shown to be comparable to traditional face-to-face methods commonly used in clinical practice, including the Mini-Mental State Examination (MMSE) [3], Repeatable Battery for the Assessment of Neuropsychological Status [4], and Montreal Cognitive Assessment (MoCA) [5]. However, these screening tools still require video administration which limits adoption in clinical practice. Also, their clinical utility is limited due to their low sensitivity [6] and lack of comparison to population averages. At best, they provide a quick snapshot of cognitive function but miss the early stages of disease and cannot specify patterns of impairment seen in various dementias. Therefore, developing a remotely accessible, self-administered, reliable, and valid CNT with higher sensitivity is needed now, more than ever.

This study used BrainCheck, a software battery capable of self-administration that provides rapid computerized neurocognitive tests that assess various areas of cognition such as attention, processing speed, coordination, memory, and executive functioning. The software has been previously validated in detecting concussion [7] and disorders of memory [8] and is classified as a diagnostic aid by the FDA. BC is already designed for remote use and is available on three different platforms: iPhone, iPad, and computer browser. In this study, we aimed to compare BC performance across multiple platforms and when administered the test vs. self-administered the test. Our goal was to validate remote self-administration of BC, particularly during the global health pandemic.

## Methods

### Participants

A mixed-method of convenience and snowball sampling was used to recruit participants that were age 50 or older and aimed to include approximately 100 participants. Internal employees of BrainCheck Inc were asked to invite volunteers and any participants were also welcomed to invite other potential participants. Enrollment and data collection occurred on a rolling basis between April 9, 2020, to May 4, 2020. Individuals that were interested completed a screening survey to capture available platforms for use: iPad, iPhone, or computer. Based on these available platforms, individuals were assigned to two possible groups.

Those assigned to Group A were to self-administer the test on at least two different BC compatible platforms: iPad, iPhone, or computer browser. The ordering of testing was left up to the participant.

Those assigned to Group B were to both self-administer and be administered the test over the phone or video. The ordering of testing was randomly assigned so that half would self-administer first and the other half would self-administer second. If a participant was exclusively assigned to Group B, they tested using the same platform. A participant was allowed to participate or 3rd or more times once their Group B participation was completed and they could then retake the test on a different platform on their own. Therefore, some participants participated in both Group A and Group B but only in the direction of Group B first then crossover to Group A.

### Procedure

114 individuals expressed interest, were assigned a group, and sent an email with instructions for completing the study. Regarding time between tests, those in Group A were instructed to wait a day between each test. For those in Group B, appointments for test administration were set up which allowed control for time between tests. All participants took BC at least twice and up to four times. Participants were given the choice of receiving their test results after completing all their tests. For those that were administered the test, the administrator and participant spoke over the phone or video and if requested, help was provided to get set-up. The administrator stayed on the phone or video for the duration of the test with the participant. It was stated the administrator was there to answer any questions that may come up during the test.

### Measurements

The CNT used was a custom BC battery of eight neurocognitive tests based on commonly included instruments in neuropsychological test batteries for the detection of cognitive impairment. Immediate and Delayed Recognition, Trail Making Test A and B, Stroop Task, Flanker Task, Coordination, and Digit Symbol Substitution Task. Additionally, the Matrix Problems Task, adapted from the Raven Standard Matrices Test, was added to the battery of assessments to measure fluid intelligence (ie, the ability to reason and problem solve), a skill that commonly declines with age [7]. The computer browser version does not include the Coordination. BC provides written instructions before each test and if applicable, practice opportunities.

For each assessment we calculated the metrics based on reaction time and correction, for example, number of correct answers to Immediate Recognition. We also calculated the composite score by combining the metrics of assessments. We then normalized these metrics against the normative population in different age groups to generate the normalized metrics.

### Analysis

For group A, one-way ANOVA with blocking was performed with platform type as a categorical factor for each assessment metrics and duration to complete the assessment. We considered each participant as a block in ANOVA. For the metrics showing significant differences on different platforms, post-hoc pairwise comparisons are performed.

For group B, two-way ANOVA with blocking was performed as well for each assessment metrics and duration to complete the assessment. We considered each participant as a block and included two categorical factors: the order the test was taken and administering status (self-administered or administered).

For all the statistical tests, p values less than 0.05 are considered a significant difference.

## Results

### Demographics

Please see Table 1 for participant demographic information.

**Table 1:**
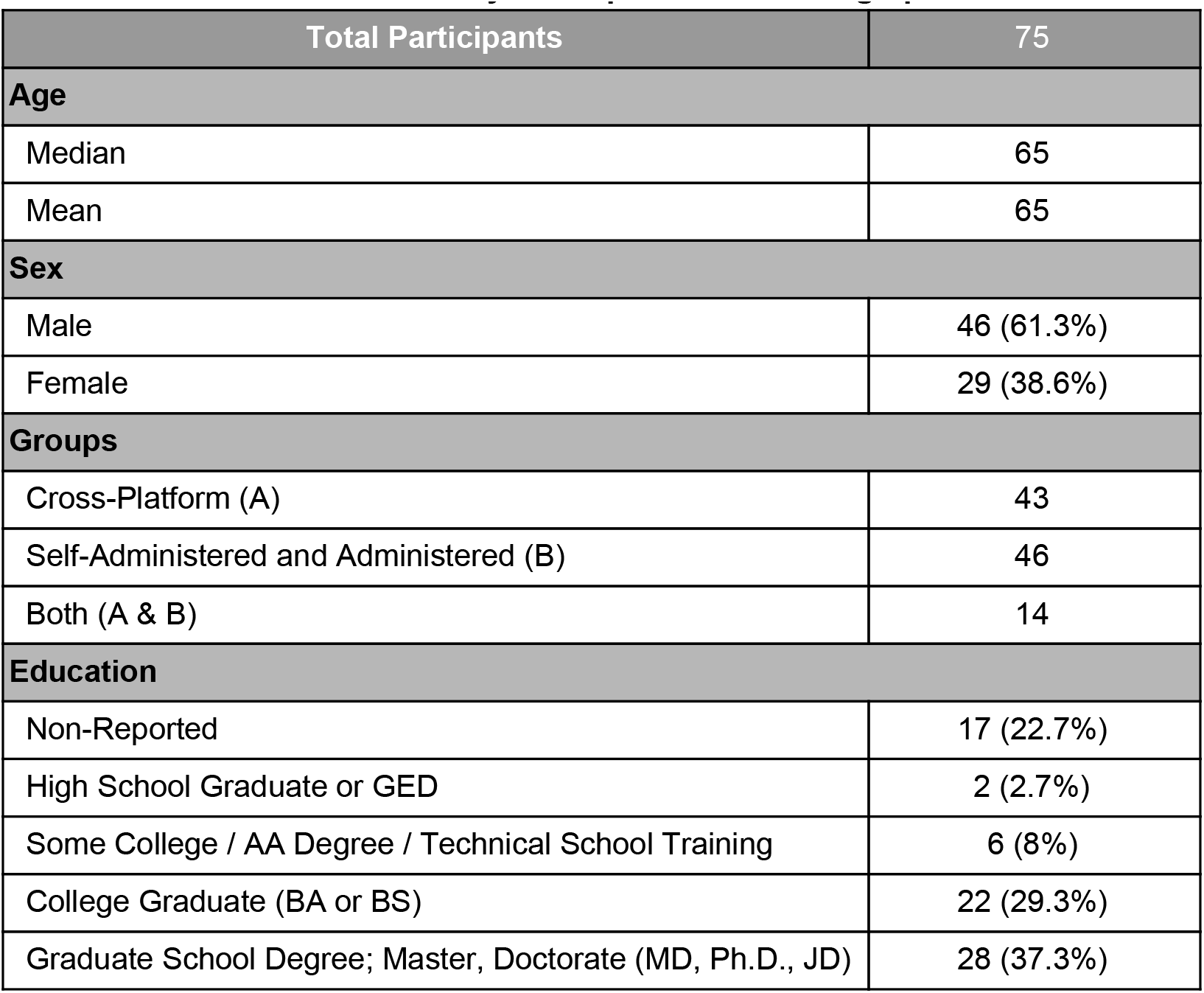
Study Participant Basic Demographics Total Participants.

| Total Participants | 75 |
| --- | --- |
| **Age** |  |
| Median | 65 |
| Mean | 65 |
| **Sex** |  |
| Male | 46 (61.3%) |
| Female | 29 (38.6%) |
| **Groups** |  |
| Cross-Platform (A) | 43 |
| Self-Administered and Administered (B) | 46 |
| Both (A & B) | 14 |
| **Education** |  |
| Non-Reported | 17 (22.7%) |
| High School Graduate or GED | 2 (2.7%) |
| Some College / AA Degree / Technical School Training | 6 (8%) |
| College Graduate (BA or BS) | 22 (29.3%) |
| Graduate School Degree; Master, Doctorate (MD, Ph.D., JD) | 28 (37.3%) |

### Cross-Platform Comparison (Group A)

A total of 99 batteries were completed, 43 on a computer browser, 30 on an iPhone, and 26 on an iPad. We found that there were significant differences in both the raw and normalized metrics across platforms for Stroop, Digit Symbol, and Trail A/B, while Flanker, Coordination, Matrix, Immediate and Delayed Recognitions had consistent performance across platforms (Table 2, Figure 1). The cross-platform difference for raw composite score did not reach significance until after normalization, which eliminates the effect of age. For the metrics showing significant differences on different platforms, post-hoc pairwise comparisons were performed between platforms. In most cases the browser differed most from iPad; specifically, the response time and duration of the assessments (Stroop, Digit Symbol, and Trail A/B) on the browser were longer than the ones on iPad.

**Table 2:**
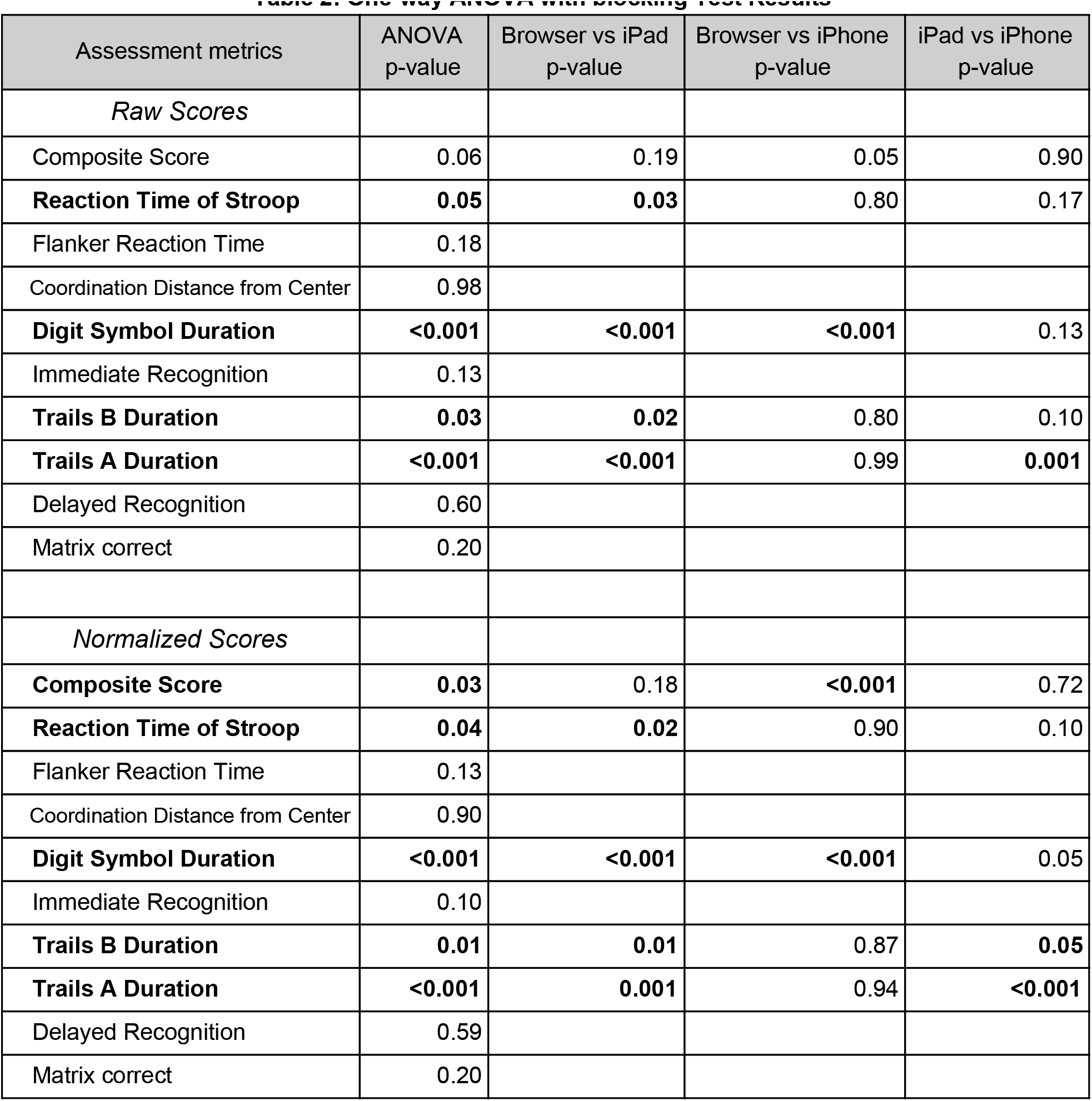
One-way ANOVA with blocking Test Results.

**Figure 1:**
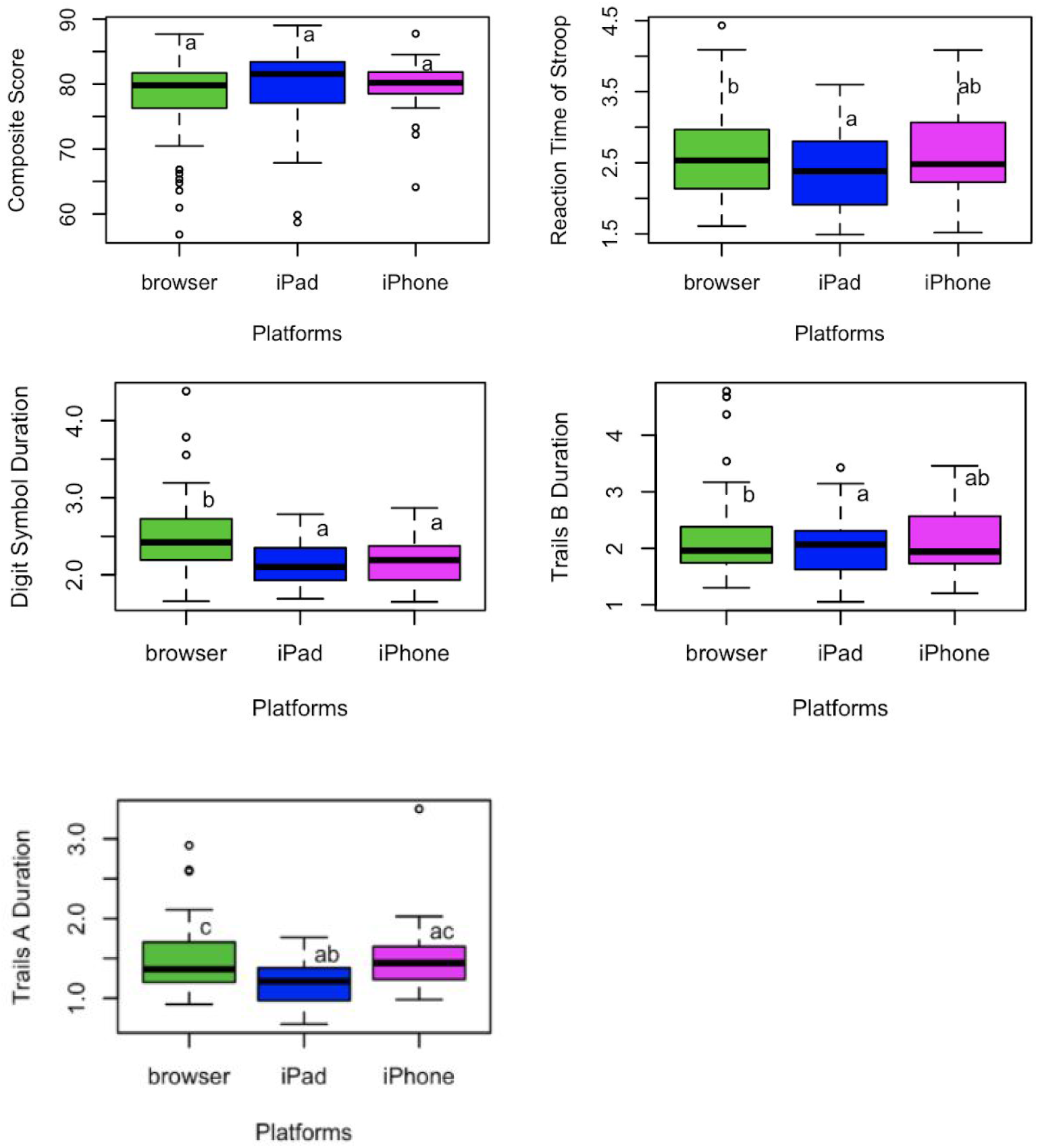
Assessments with Significant Different Metrics Across Platforms. Raw metrics of the assessments showing significant differences are shown. Different letters indicate significant differences.

In addition, we compared the time needed to complete the battery on different platforms. We found that the duration of completing a battery was significantly different among platforms (p<0.001). The post hoc analysis showed the participants who completed BC on iPhone took less time compared to both iPad (p=0.039) and computer browser (p<0.001) but there was no difference between using computer browser and iPad (p=0.91) (Figure 2).

**Figure 2:**
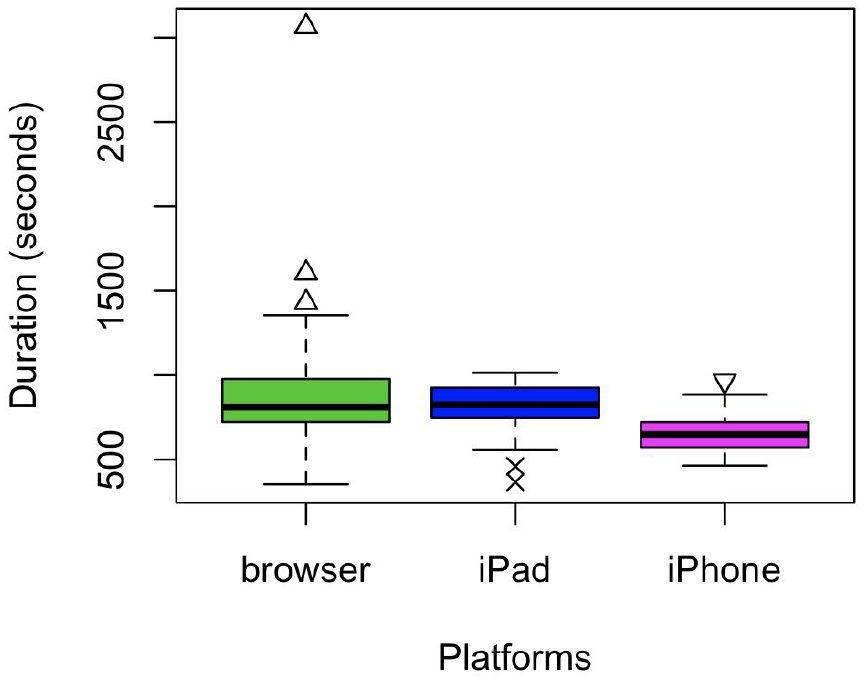
Comparison of Duration to Complete BC in Group A.

### Self-Administered and Administered Comparison (Group B)

In the administration comparison group, 46 participants took both self-administered and administered tests in a random order. 30 participants took self-administered tests first and 16 took administered tests first. We used ANOVA by considering both administering status and the order of tests (i.e. first/second test) as categorical factors and each participant as the block. We found no significant impact on the results between administered and self-administered tests except for the Coordination test, which is mainly due to lack of data. However, the order of tests taken did affect some assessments significantly, which include Stroop, Flanker, and Digit Symbol (Table 3). A closer look at these significant assessments (Stroop, Flanker, and Digit Symbol) shows that response time was shorter in the second test (Figure 3), which indicates the participants may have become familiar with the assessments after the first test.

**Table 3:** Two-way ANOVA with blocking Test Results.

| Test metrics | Order of test | Administering status |
| --- | --- | --- |
| *Raw Scores* |  |  |
| Composite Score | 0.92 | 0.83 |
| **Reaction Time of Stroop** | **0.04** | 0.12 |
| **Flanker Reaction Time** | **<0.001** | 0.35 |
| **Coordination Distance from Center** | 0.05 | **0.02** |
| **Digit Symbol Duration** | **<0.001** | 0.75 |
| Immediate Recognition | 0.77 | 0.35 |
| Trails B Duration | 0.59 | 0.18 |
| Trails A Duration | 0.44 | 0.29 |
| Delayed Recognition | 0.72 | 0.67 |
| Matrix Correct | 0.72 | 0.38 |
| *Normalized Scores* |  |  |
| Composite Score | 0.96 | 0.86 |
| **Reaction Time of Stroop** | **0.04** | 0.14 |
| **Flanker Reaction Time** | **0.002** | 0.46 |
| **Coordination Distance from Center** | 0.05 | **0.02** |
| **Digit Symbol Duration** | **< 0.001** | 0.94 |
| Immediate Recognition | 0.67 | 0.43 |
| Trails B Duration | 0.57 | 0.20 |
| Trials A Duration | 0.50 | 0.26 |
| Delayed Recognition | 0.48 | 0.73 |
| Matrix Correct | 0.76 | 0.41 |

**Figure 3:**
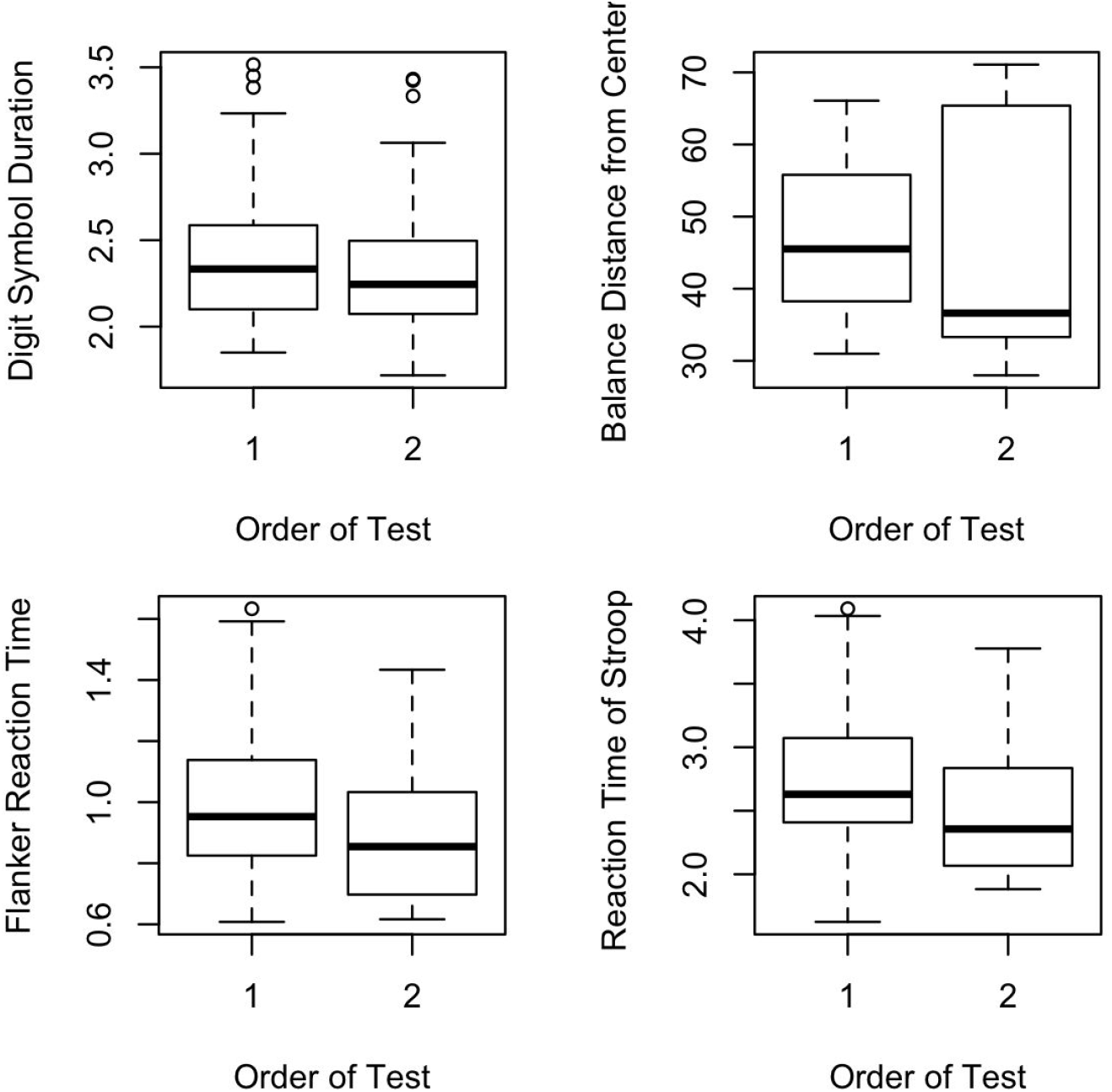
Significantly Different Assessment Metrics Between the First and Second Tests.

In addition, we compared the time needed to complete the battery at different administration types and the order of tests. We found that there is no significant difference in duration to complete the battery for either factor regardless of platform (Figure 4).

**Figure 4:**
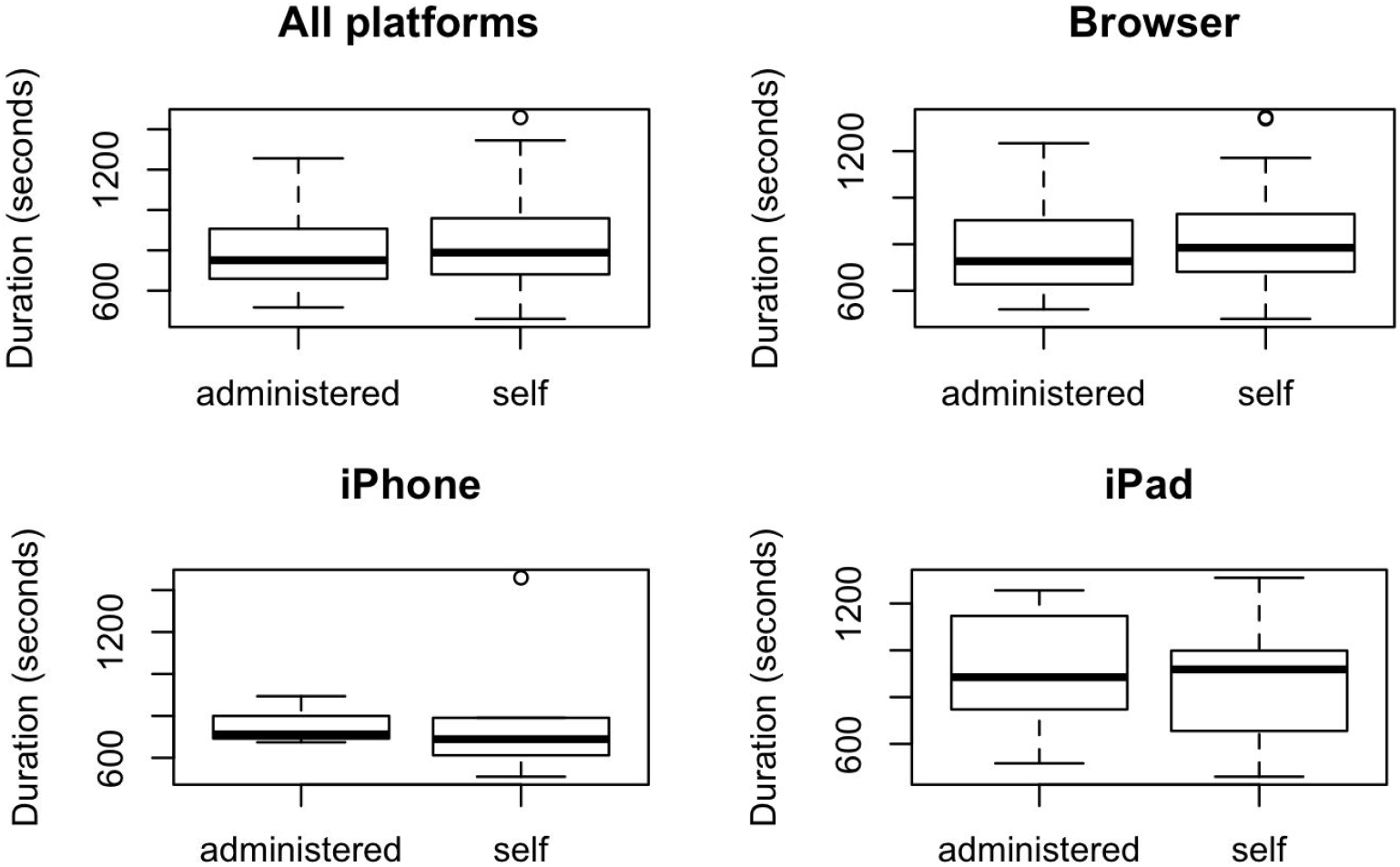
Comparison of Duration to Complete BC under Different Administration Types.

## Discussion

In this study we evaluated the reliability and utility of BC used remotely on different platforms and with and without administration by enrolling 75 participants to take BC in different settings. Overall BC test results were consistent whether administered or self-administered, while significant differences in performance were found regarding platform use. These results indicate that BC could be a reliable and self-administrated tool to assist remote cognitive testing with an adjustment of platforms.

In cross-platform comparison we found Stroop, Digit Symbol, and Trail A/B had significantly different performance across the platforms, while Flanker, Coordination, Matrix, Immediate and Delayed Recognitions did not. For Stroop, Digit Symbol, and Trail A/B assessments, this is likely due to innate differences of the interactions between the platforms. All the mentioned assessments require the participants to locate and click the correct answers on the screen to complete the task. The participants used a mouse or trackpad on the computer browser version, while on iPad and iPhone they could touch the screen directly to complete the task. This may explain the consistent trend in response time and duration when we compared these assessments performance on the browser to the iPad. For the Flanker assessment participants used touch screens to complete the tests on iPad or iPhone and they used a keyboard (only q and p letter keys) to complete the computer browser version of the assessment. The usage of the keyboard likely reduced the reaction time to a similar level of using the touch screens on iPad or iPhone. The Coordination assessment is only available on iPad or iPhone, which are very similar platforms. For the Matrix and Immediate and Delayed Recognitions assessments, the metrics we used are the number of correct responses to the assessment, which does not depend on the reaction time or duration. This may explain why their performances are consistent across platforms. While we considered looking at the effects from order of test taking for cross platform use, not all participants contributed data for each platform, and there was a bias toward taking the test first on a browser, making it difficult to truly evaluate the effect of order. Overall, these findings indicated that it would be necessary to adjust for platform types in assessment scoring, which is currently the standard in the BrainCheck scoring algorithm. This reinforces our previous findings and current methods of standardizing the composite score by platform type.

Within Group B, we saw that the type of administration had no impact on test results. Again, while the BC composite score was similar in the two tests taken by a participant, we found whether it was the first time or the second time the test was taken mattered for three of the assessments, impacting a total of 6 of 20 assessment metrics measured. These include metrics within the Flanker, Stroop, and Digit Symbol tests (Figure 3). These results likely indicate the existence of the practice effects on certain tests when repeated in close proximity. To further check if this effect faded away as the interval between two tests increased, we checked the dependence of difference between the two tests on the interval time by categorizing those completed on the same day and those completed on two separate days. However, we did not detect any significant differences which indicates that such practice effects did not change over time within our study time frame. It is possible that if we get more data over longer periods, there could be a trend that the differences in the two tests diminishes over time. It is notable that in clinical practice this issue is unlikely to be present given in a typical patient scenario BC testing would not happen within the same day and unlikely even within the same week or month.

It is also important to understand any factors that may impact the validity of these remote neurocognitive testing. A major weakness of this initial work is more than 90% of the participants have college degrees. Future research will need to involve more people with diverse education levels and check impacts of such factors. Additionally, the sample size was relatively small and unbalanced, owing to the compressed timeline due to the global health pandemic. In the cross-platform comparison, not every participant completed the tests on all three platforms. Also, we observed the sequence of platforms for each participant was not totally random where most participants completed their first test on the browser. Furthermore, there are a number of factors at play here that were not controlled for including the impact of having environmental noise and conditions. We aimed to allow for a natural environment to capture a typical remote testing experience. There may be factors for some patients in their home environments which may directly impact their test scores. We also did not differentiate computer browsers based on interface such as if a mouse or a trackpad was present, which may impact performance. All these ought to be considered in future testing opportunities.

## Conclusion

In our current state, there is an unprecedented need for tele-medicine, –health, and –testing. These findings show the fairly consistent testing capabilities available now when delivering remote neurocognitive testing using BrainCheck in a variety of administration methods. Both self-administered and administered tests demonstrate consistent results providing initial validation for at home testing. To address the differences seen across platforms we currently provide standardized scores by platform. Given these findings and considerations, BrainCheck may be used remotely to reach patients at home via telehealth.

## Data Availability

Data may be made available by contacting the corresponding author and with a data use agreement

## Ethical Approval

This study protocol was reviewed and approved by Solutions IRB. All participants provided informed consent for being in the study.

## Acknowledgments

Funding was provided by BrainCheck, Inc.

## Data Availability

Data may be made available by contacting the corresponding author and with a data use agreement

